# Data-driven research on eczema: systematic characterization of the field and recommendations for the future

**DOI:** 10.1101/2022.01.14.22269294

**Authors:** A. Duverdier, A. Custovic, R.J. Tanaka

## Abstract

**Background:** The past decade has seen a substantial rise in the employment of modern data-driven methods to study atopic dermatitis (AD) / eczema.

**Objective:** To summarise the past and future of data-driven AD research, and identify areas in the field that would benefit from the application of these methods.

**Methods:** We retrieved the publications that applied multivariate statistics (MS), artificial intelligence (AI, including machine learning-ML), and Bayesian statistics (BS) to AD and eczema research from the SCOPUS database over the last 50 years. We conducted a bibliometric analysis to highlight the publication trends and conceptual knowledge structure of the field, and applied topic modelling to retrieve the key topics in the literature.

**Results:** Five key themes of data-driven research on AD and eczema were identified: (1) allergic co-morbidities, (2) image analysis and classification, (3) disaggregation, (4) quality of life, and (5) risk factors and prevalence. ML&AI methods mapped to studies investigating quality of life, prevalence, risk factors, allergic co-morbidities and disaggregation of AD/eczema, but seldom in studies of therapies. MS was employed evenly between the topics, particularly in studies on risk factors and prevalence. BS was focused on three key topics: treatment, risk factors and allergy. The use of AD or eczema terms was not uniform, with studies applying ML&AI methods using the term eczema more often. Within MS, papers using cluster and factor analysis were often only identified with the term AD. In contrast, those using logistic regression and latent class/transition models were “eczema” papers.

**Conclusions:** Research areas that could benefit from the application of data-driven methods include the study of the pathogenesis of the condition and related risk factors, its disaggregation into validated subtypes, and personalized severity management and prognosis. We highlight Bayesian statistics as a new and promising approach in AD and eczema research.

## INTRODUCTION

Atopic dermatitis (AD, also referred to as eczema or atopic eczema) is a common chronic inflammatory skin disease that affects approximately 20% of children and 10% of adults in high-income countries^1^. Recently, computational modelling^2^ and data-driven analytical methods have emerged as powerful new approaches to AD research, especially to elucidate its complex pathophysiology^3^, patient-dependent response to treatment^4,5^, and endotypes or subtypes^6–11^.

Big data have revolutionized the way we study disease^12^. The increased availability of large and diverse medical datasets has favoured the adoption of modern computational methods which can integrate and interrogate large quantities of data and extract hidden patterns and associations. There are three primary analytic methodologies or disciplines for data-driven research: multivariate statistics (MS), Bayesian statistics (BS), and machine learning and other artificial intelligence methods (ML&AI). MS encompasses methods that analyse datasets with multiple independent and/or dependent variables^13^, which is a key characteristic of biomedical datasets thereby making MS a popular and powerful methodology. AI is a field concerned with building systems that can mimic human intelligence, and ML is a subfield of AI. Finally, BS allows us to combine prior knowledge and observed data^14^, contrasting the frequentist approach which bases its analysis only on the observed data^12,15^, and is a potentially promising approach to develop predictive models and utilize clinical data. Such data-driven approaches have been applied to identify biomarkers to diagnose disease and identify therapeutic targets^12,16,17^. Deep neural networks have been developed to aid in the detection and diagnosis of skin^18^, breast^19^, and prostate^20^ cancer. In AD research, the Bayesian mechanistic model recently developed by Hurault et al.^21^ can predict individual patients’ next-day AD severity scores from their previous severity scores and treatments applied. These examples illustrate the benefit of employing a data-driven approach in medical fields with a growing quantity of data.

Within the AD community, data collection is increasing, providing a unique opportunity to leverage data-driven methods^2^. As we enter a period of further substantial growth in the employment of data-driven methods to study AD, we aimed to identify the areas in AD research where data-driven methods have been applied, their current state of development, and highlight the knowledge gaps in the field that could benefit from the application of these methods. To address our aim, we conducted a bibliometric analysis highlighting the publication trends and conceptual knowledge structure of data-driven research on AD and eczema, and applied topic modelling to retrieve the key topics present within the literature. Bibliometrics uses statistical tools to study publication trends and patterns within an area of research^22,23^, and can be used to summarise a field of research in a systematic and reproducible manner. Probabilistic topic modelling explores the knowledge structure of a field by identifying the latent thematic structure of a corpus of documents^24^. A bibliometric analysis was previously conducted to understand the knowledge structure and theme trends of AD research^25^ but it considered publications with the term AD from 2015 to 2019 and did not focus on data-driven research. Also of note, the continued absence of a consensus in nomenclature has resulted in the co-existence of two main terms for the skin condition, AD and eczema, which have been shown to be linked to different findings and biased to different disciplines^26^. Our study included both AD and eczema terms and retrieved all publications available up to March 2021 without a time constraint, to provide the full picture of the field. Additionally, we included topic modelling to provide a detailed view of the key research topics in the field and methods employed.

## METHODS

This section summarises the analysis conducted in this paper; detailed description is presented in Appendix S1.

### Literature Search

We retrieved all publications to March 17^th^, 2021, on AD and eczema that apply MS, ML&AI, and BS methodologies from the SCOPUS database. The keywords ‘atopic dermatitis’ and ‘eczema’ were used with each of MS, ML&AI, and BS methodologies.

### Bibliometric Analysis

We performed a bibliometric analysis on the bibliographic information (including the authors, sources, countries, citations, and keywords) of the publications obtained from the literature search. Using the *bibliometrix* R package^27^, we obtained descriptive statistics on the collection of publications, including the most productive countries and the general publication trends. We also performed co-word analysis to produce keyword co-occurrence networks and thematic maps.

### Probabilistic Topic Modelling

We used the Latent Dirichlet Allocation (LDA) algorithm^28^ to explore the main topics present in the publications obtained by the literature search. LDA is an unsupervised ML method that estimates both the distribution of topics within each document and the distribution of words within each topic, by assuming each document consists of a mixture of topics and each topic consists of a mixture of words. Here, each document consisted of the title, keywords, and abstract. We used the *tm* R package^29^ to clean the data (tokenization, lowercase conversion, removal of special characters and stop-words, standardization of words) and remove words with low frequency (words that occurred in less than ten publications), the *topicmodels* R package^30^ to run the LDA algorithm on the corpus, and generated the plots of the results using R packages such as *ggplot2*^31^ and *wordcloud*^32^.

## RESULTS

### Publication trends of data-driven research on AD and eczema

620 unique articles, published between September 1973 and March 2021, were retrieved from the SCOPUS database on the application of MS, ML&AI and BS to AD/eczema research. Of these, 474 articles employed MS methods, 150 ML&AI, and 37 BS (Table S1, Figure S1). Publications increased over time, with most papers published in the past decade (473/620, 76.3%) (Figure 1). The earliest application of MS in AD and eczema research dates to 1973, ML&AI made its first appearance in 1996, and BS in 2001 (Figure 1A). The application of ML&AI has seen a particular emergence in the past ten years, with a substantial increase since 2019, approaching the popularity of MS. BS is the least commonly used of the three methodologies to date. More details on publication trends are presented in Appendix S2.

**Figure 1:**
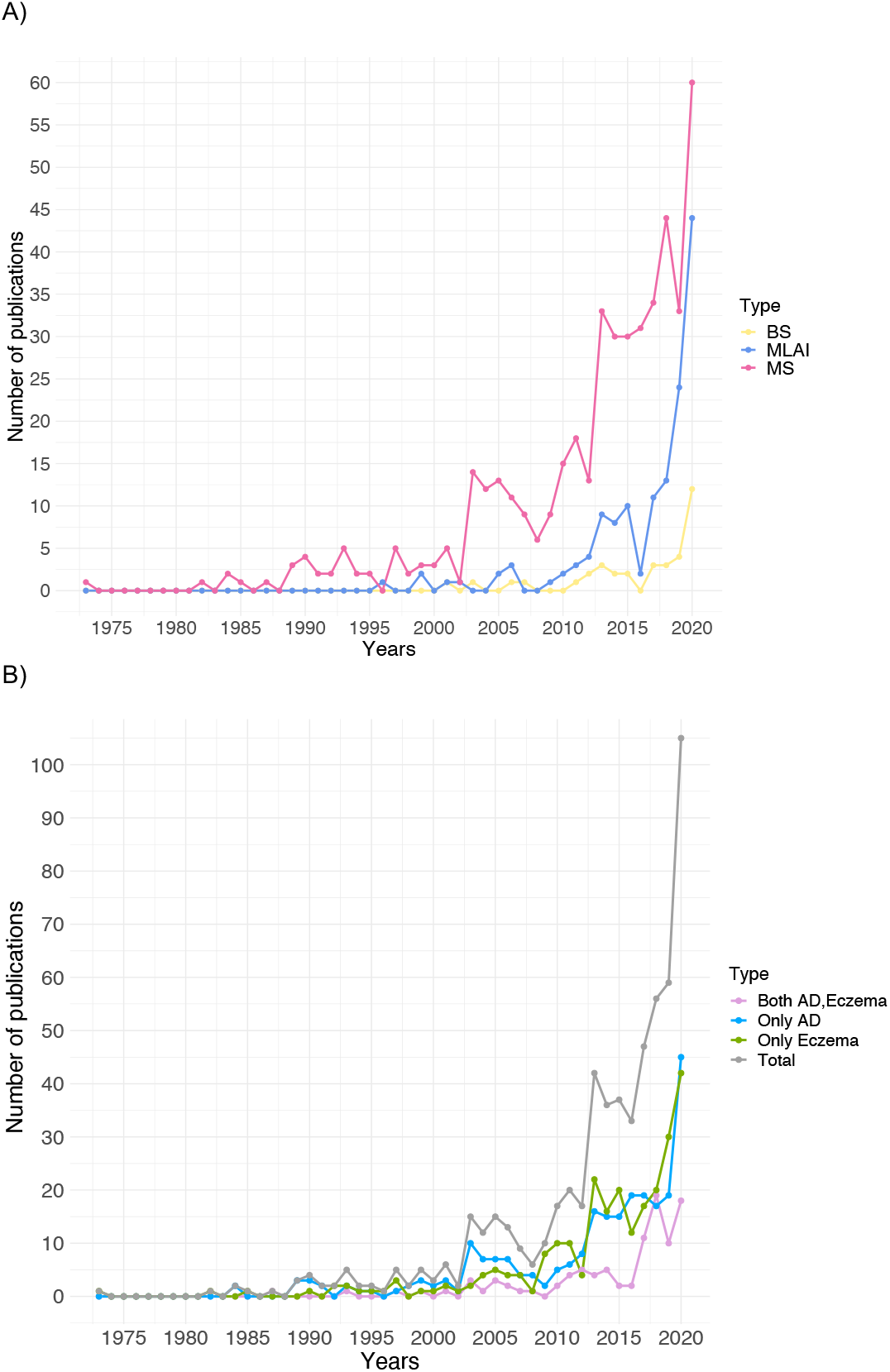
Papers published per year separated by **(A)** methodology and **(B)** term.

Most publications are labelled as either AD or eczema, and only a small portion are annotated with both terms (100 of 620 articles). This phenomenon is similarly found within the individual methodologies (Table S1). Publication numbers for each term are similar throughout the years, showing at first glance no significant frequency preference of the field in general for one term over the other (Figure 1B). Geographical distribution of manuscripts is shown in Table S2.

### Analytical methods and the use of AD or eczema terms

Table 1 summarizes the key methods used within the collection of publications. Cluster and factor analysis are the two most common methods. Of the 37 BS papers, a manual inspection found that only eight^21,33–39^ specifically study AD. Of these eight, half^33–36^ used random-effects Bayesian network meta-analysis to compare treatments for AD, and one^21^ uses a Bayesian mechanistic machine learning model to predict next-day AD severity for individual patients.

**Table 1.** Distribution of the main methodologies used in data-driven eczema and atopic dermatitis (AD) publications. Search strings representing the methodologies were searched for in the title, abstract, and keywords of the multivariate statistics (MS) and machine learning and artificial intelligence (ML&AI) publications. Methods for Bayesian statistics (BS) were determined manually. Number of publications are given for the total collection and additionally separated according to the term used, only eczema, only AD, or both. For each method, the highest number of publications between only eczema and only AD is bolded.

| Discipline | Methodology | Number of Publications |  |  |  |
| --- | --- | --- | --- | --- | --- |
|  |  | Total<br>(of 620) | Only<br>Eczema<br>(of 255) | Only<br>AD<br>(of 265) | Both<br>(of 100) |
| Multivariate<br>statistics<br>(MS) | Cluster analysis | 206 | 84 | **90** | 32 |
|  | Factor analysis | 89 | 26 | **46** | 17 |
|  | Logistic regression | 56 | **24** | 17 | 15 |
|  | Latent class/Transition | 55 | **31** | 12 | 12 |
|  | Principal component analysis | 46 | 17 | **20** | 9 |
|  | Discriminant analysis | 28 | 8 | **16** | 4 |
|  | Markov model | 28 | 8 | **12** | 8 |
|  | Structural equation modelling | 11 | **5** | 4 | 2 |
|  | Mixture model | 7 | 3 | **4** | 0 |
|  | Correspondence analysis | 3 | **2** | 1 | 0 |
|  | Latent variable model | 3 | **2** | 1 | 0 |
|  | Canonical correlation | 1 | 0 | **1** | 0 |
| Machine<br>learning and<br>artificial<br>intelligence<br>(ML&AI) | Artificial neural networks | 67 | **44** | 21 | 2 |
|  | Machine learning | 48 | **25** | 17 | 6 |
|  | Support vector machine | 36 | **24** | 11 | 1 |
|  | Artificial intelligence | 17 | **8** | 7 | 2 |
|  | Decision trees | 17 | **7** | 6 | 4 |
|  | Deep learning | 13 | **9** | 3 | 1 |
|  | Natural language processing | 12 | **7** | 5 | 0 |
|  | Random forests | 9 | **6** | 2 | 1 |
|  | Supervised learning | 2 | **2** | 0 | 0 |
|  | Unsupervised learning | 1 | **1** | 0 | 0 |
| Bayesian<br>statistics<br>(BS) | Bayesian framework | 14 | **8** | 5 | 1 |
|  | Bayesian network | 5 | **4** | 1 | 0 |
|  | Random-effects Bayesian | 4 | 0 | 1 | 3 |
|  | Bayesian machine learning | 3 | **2** | 0 | 1 |
|  | Bayesian spatial and temporal | 3 | 1 | **2** | 0 |
|  | Naïve Bayesian classifier | 2 | **2** | 0 | 0 |
|  | Bayesian meta-regression | 2 | 0 | 1 | 1 |
|  | Bayesian model averaging | 2 | **2** | 0 | 0 |
|  | Bayesian latent class analysis | 1 | **1** | 0 | 0 |
|  | Random-effects Bayesian | 1 | 0 | **1** | 0 |

The use of AD and/or eczema terms is not uniform throughout the different methods. Detailed analysis is presented in the Appendix S3. Briefly, papers applying ML&AI methods use the term eczema more often. Within MS, papers using cluster and factor analysis are often only identified with the term AD. In contrast, those using logistic regression and latent class/transition models are eczema papers.

### Five central themes of data-driven AD and eczema research and their level of development

The bibliometric analysis identified five key themes within AD/eczema research employing MS, BS, and ML&AI methods, as visualized in a thematic map (Figure 2), where themes are mapped onto a two-dimensional space according to their centrality and density. The centrality is the degree of interaction of the theme with other themes and measures the significance or relevance of a theme in the development of the field at large^40^. The density measures the development of the theme^40^. Using these two measures, themes can be separated into four quadrants: emerging or declining themes (low centrality and density), niche themes (low centrality and high density), motor themes (high centrality and density), and basic themes (high centrality and low density)^40^. We named the five identified themes retroactively, ordered by decreasing density:

**Figure 2:**
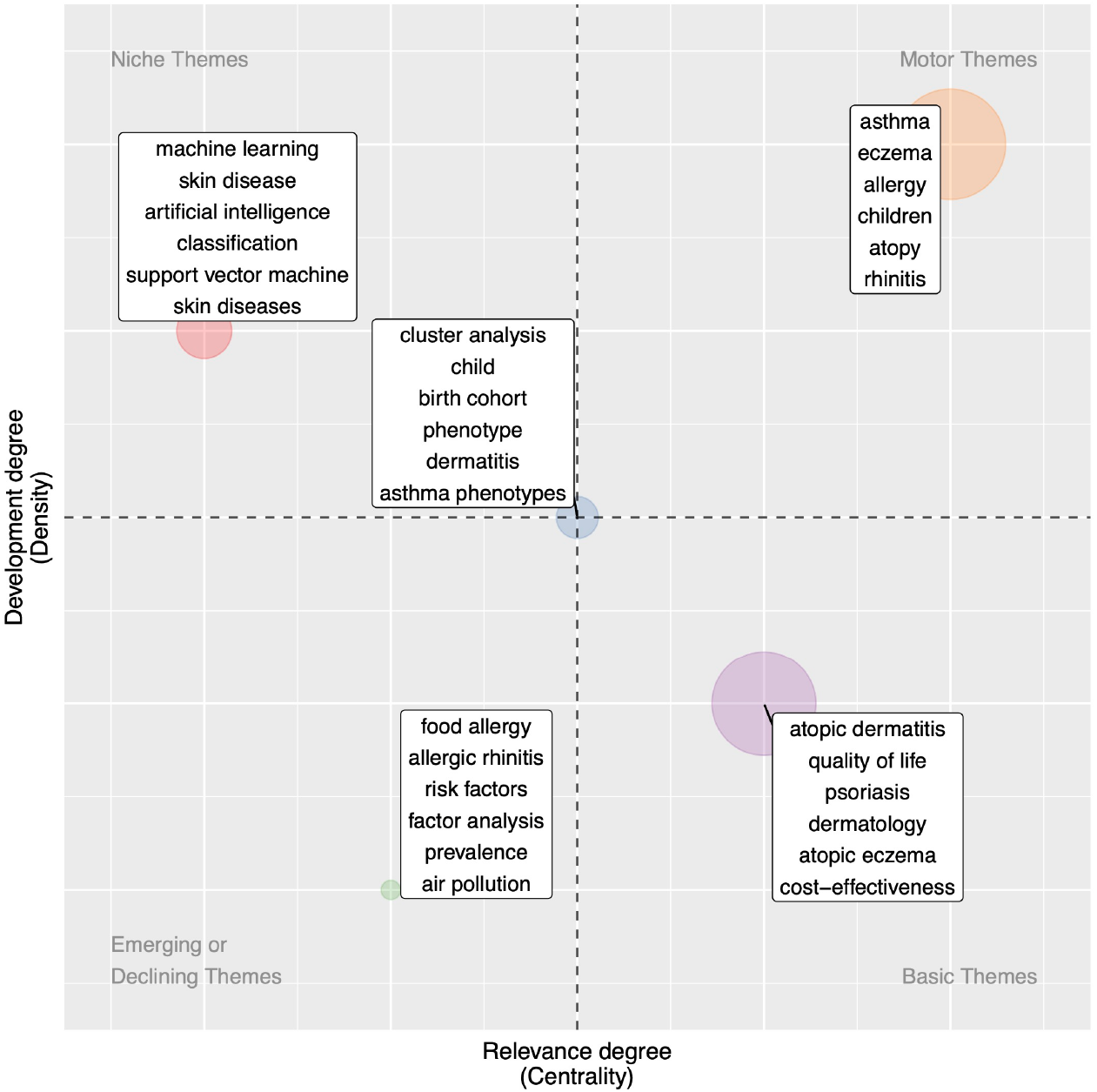
Thematic map. Themes were generated using the top 100 authors’ keywords and separated according to centrality (the degree of interaction of the theme with other themes) and density (the strength of internal connections among keywords in the theme). Up to six of the most frequent keywords in the associated theme are shown on the map.

<u>Theme 1: Allergic co-morbidities</u>. This theme includes articles that study atopic/allergic diseases and the development of IgE sensitisation in childhood using analytical methods such as cluster analysis, latent class analysis, and AI. It is a motor theme, highly relevant and already developed (orange in Figure 2).

<u>Theme 2: Image analysis and classification.</u> This theme includes articles that use ML methods, including deep learning and support vector machines, to process, segment, and classify images of AD/eczema and other skin diseases. It is a niche theme with a high development degree but is less central than other themes in the field (red in Figure 2).

<u>Theme 3: Disaggregation of the condition.</u> This theme tackles the issue of disentangling the complex pathophysiology of AD/eczema and includes studies that consider biomarkers to investigate endotypes and methods such as cluster analysis to disaggregate phenotypes. The theme also contains articles on disaggregating asthma phenotypes. Compared to other themes, it has middle relevance and development (blue in Figure 2).

<u>Theme 4: Quality of life.</u> This theme includes studies investigating the quality of life and the cost-effectiveness of treatments, not specific to only AD/eczema but also for other skin conditions such as psoriasis. It is a basic theme with relatively low development but high relevance (purple in Figure 2).

<u>Theme 5: Risk factors and prevalence.</u> This theme looks at potential factors that increase the likelihood or prevalence of AD/eczema, including air pollution. It also studies the relationship with co-morbidities, including allergic rhinitis and food allergy. Methods used in this theme include factor analysis. It is an emerging theme with low development and relevance (green in Figure 2).

Thematic maps were also generated for the three methodologies and the term used (eczema or AD), Figure S2.

### Eight key topics and identified gaps in employing modern computational methodologies

The LDA algorithm revealed eight key topics of data-driven AD and eczema research (Figure 3, Table S3), by breaking down the five themes obtained in the bibliometric analysis into their main components. It identified, in greater detail, the key areas of interest explored in the literature to date (Table S4) and their growth over time (Figure S3).

**Figure 3:**
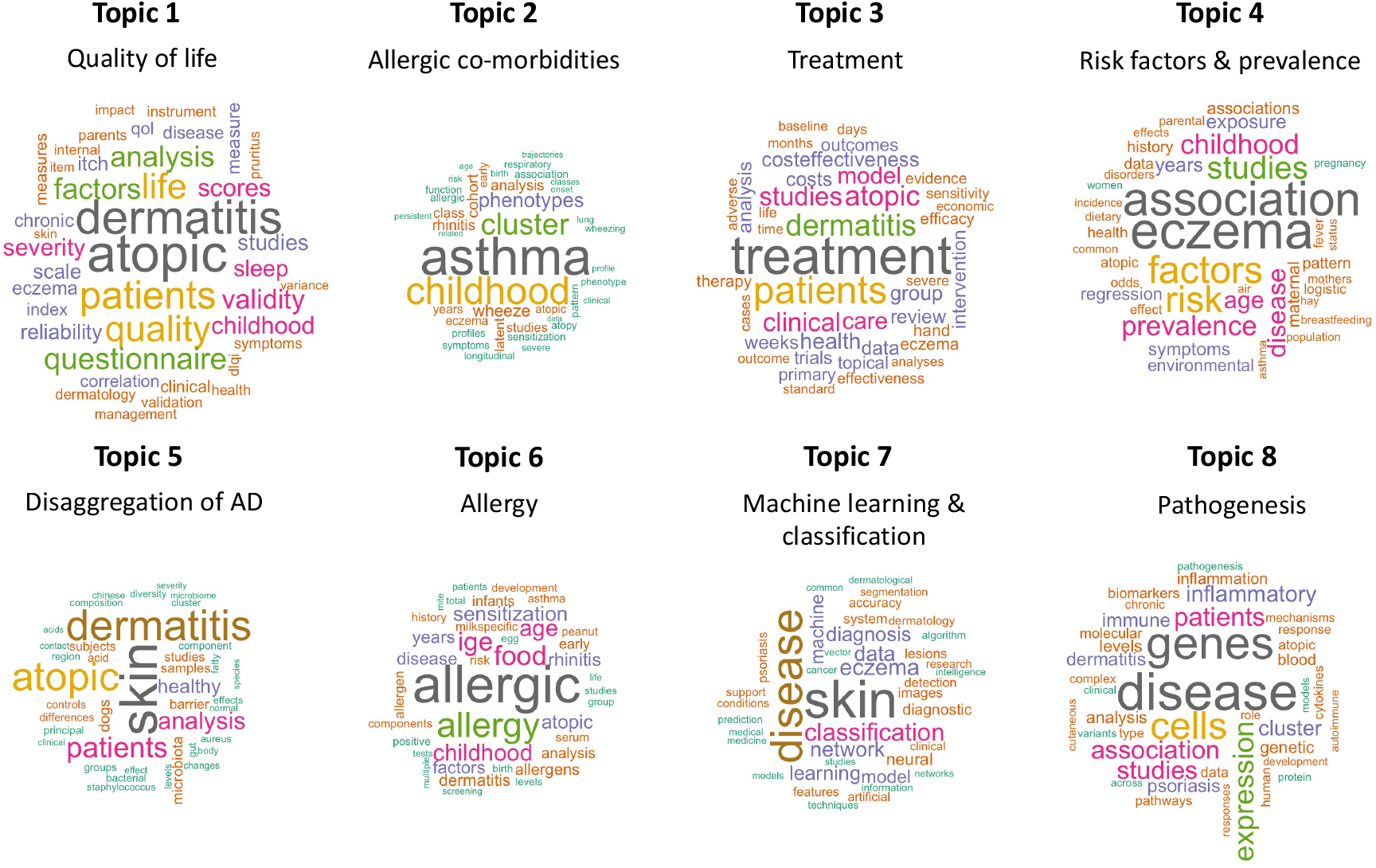
Word clouds for the eight topics obtained by Latent Dirichlet Allocation (LDA). The topic names were retroactively chosen to best summarize the content of topics. The 40 most probable words in each topic are plotted with the size of the words proportional to their probability.

Theme 1 is represented by Topics 2 and 6, which respectively study allergic co-morbidities and the development of allergy and sensitisation. Theme 2 is represented in Topic 7 on the application of ML methods for the classification and diagnosis of skin diseases. Theme 3 is broken down into two key topics, Topics 5 and 8. Topic 5 includes studies on the disaggregation of AD and the role of the skin microbiome, especially the presence of *Staphylococcus aureus*. Topic 8 regards the pathogenesis of the condition, looking at genetic, inflammatory and immune biomarkers, and mechanisms underlying development and progression. Theme 4 encompasses Topics 1 and 3, which study AD/eczema symptoms and their management. Topic 1 includes publications on disease severity and the effect on quality of life. Topic 3 encompasses therapeutic studies, including efficacy and cost-effectiveness analyses. Finally, Theme 5 is reflected in Topic 4, which considers the prevalence of eczema and associated risk factors, including environmental exposure and parental history of atopy.

Most publications that employ ML&AI are found in Topic 7 (Figure 4). In contrast, topics including quality of life, disaggregation of AD, risk factors and prevalence, allergic co-morbidities, and treatment see a low application of ML&AI methods.

**Figure 4:**
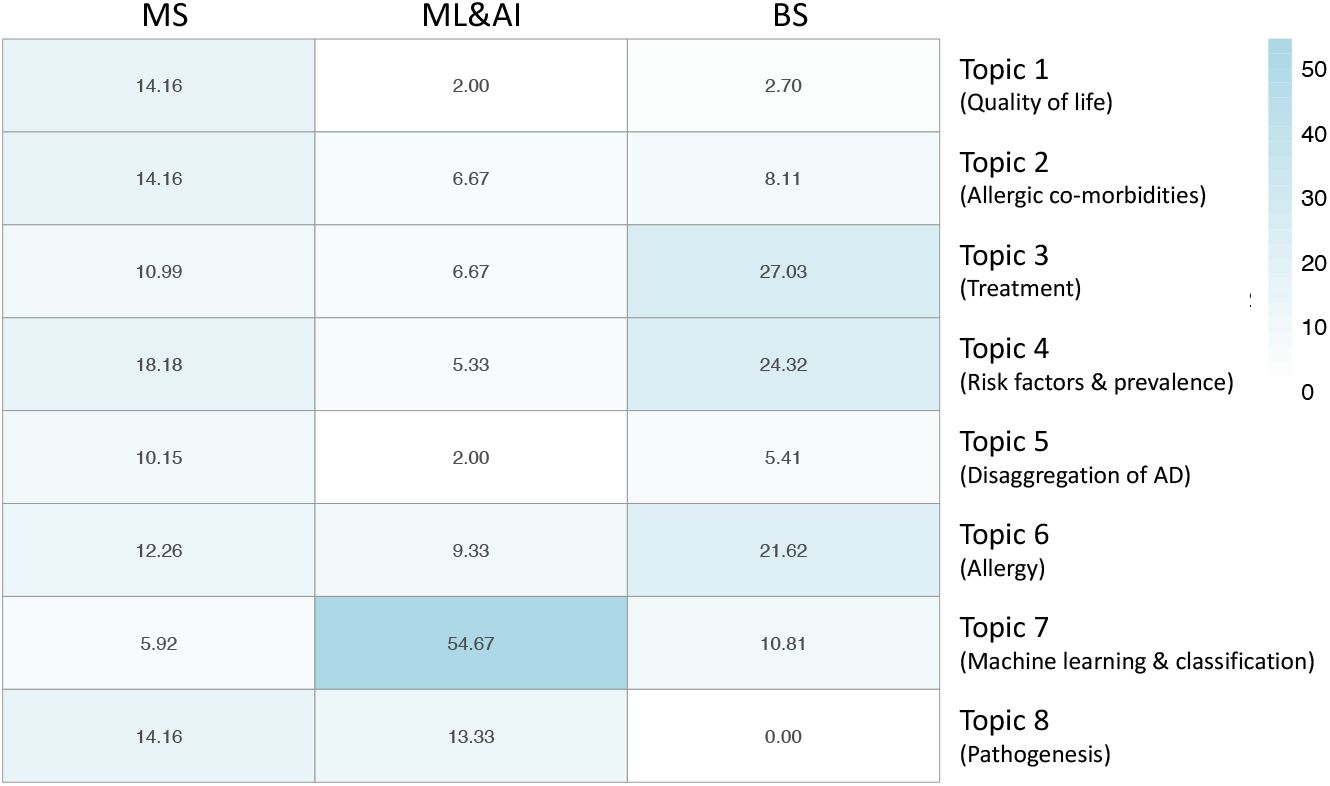
Distribution of the application of MS, ML&AI, and BS methodologies in the eight LDA topics. Each publication is assigned to its most probable topic.

MS is employed fairly evenly between the topics. Its most popular topic is Topic 4 on risk factors and prevalence.

BS focuses on three key topics: treatment, risk factors and prevalence, and allergy.

Topics on quality of life, treatment, and disaggregation of the condition contain ‘atopic’ and ‘dermatitis’ as two of the most relevant keywords, while ‘eczema’ is found as the top keyword in studies on risk factors and prevalence (Figures 4 and S4). This reflects the trend seen in the bibliometric analysis that the term eczema tends to be used in publications that also study other allergic diseases (Figure S5). In contrast, AD term is used often in publications that are more specific to the condition.

## DISCUSSION

The first application of data-driven methods to AD and eczema research occurred in September 1973. Since then, 620 articles have been published, with over three-fourths of the publications in the last decade. The growth in scientific production over time shows an increased interest in applying data-driven methodologies to the study of AD and eczema, similar to asthma research^12,15^.

Five central themes currently characterize the field: (1) allergic co-morbidities, (2) image analysis and classification, (3) disaggregation of the condition, (4) quality of life, and (5) risk factors and prevalence. In 2020, Theme 2 was the most popular topic in AD and eczema research today. Theme 3 has the third-highest number of publications, indicating the continued need to delineate developmental trajectories and disease mechanisms and the subsequent characterization and validation of endotypes before these can be used and implemented in other areas of research. Theme 4 is central to the field, but not highly researched to date, suggesting the continued need to apply data-driven methods to help build personalized severity prediction tools and treatment strategies. Theme 5 has the lowest number of publications; as more data is collected on potential risk factors, data-driven tools could be leveraged to evaluate their relevance. Mu et al.^25^ conducted a bibliometric analysis on AD literature from 2015 to 2019 and similarly found that themes on quality of life, prevention and control, and pathology were undeveloped. This reinforces the need for further research, particularly employing data-driven methodologies, in these key areas.

We identified a substantial increase in ML&AI publications over the past five years in AD and eczema research. Most studies that apply ML in dermatology address classifying skin lesions and primarily rely on convolutional neural networks for image recognition and classification^41^. Over half of the AD and eczema publications employing ML&AI methods are found in Theme 2 on image analysis and classification. The low application of ML&AI in the rest of the themes, including the study of the pathogenesis of the condition, disaggregation, risk factors, quality of life, treatment, and the role of allergic co-morbidities, highlights potential areas that could benefit from the application of ML&AI methods.

Bayesian approaches have been used to study asthma^12,15^ and the relationship between allergic diseases^42,43^. However, only eight publications to date apply BS to study AD and eczema specifically. One of the eight developed a Bayesian mechanistic model that can predict next-day AD severity from patients’ previous severity scores and treatments applied^21^. The Bayesian approach allows consistent quantification of uncertainty within parameter estimates and predictions and the incorporation of prior knowledge or data from previous studies. This highlights BS as a new and promising approach in AD and eczema research, particularly to develop predictive models and use clinical data.

The analysis performed in this study corroborates the discrepancy in the use of AD and eczema terms within the literature that has been highlighted in previous studies^26,44^. Our results point towards a bias in term use depending on the computational method employed; this alludes to the previously articulated notion that AD and eczema terms may be associated to different research communities that have differing views on nomenclature (Appendix S3).

The main limitation of our analysis is that it is heavily dependent on the terminology used by the authors. The authors’ keywords associated to each publication were used to discover the key themes of the field of research; they were also used, in part, to retrieve publications of interest. This points to the importance of keyword choice when publishing a paper and the impact of using eczema or AD terms. A second limitation is that the LDA algorithm was applied on each publication’s title, keywords, and abstract, but not the full text as they were not available. Additionally, the publications were retrieved from the SCOPUS database. Although similar in content to that found on PubMed, future systematic reviews could aggregate the publications from multiple databases to ensure completeness of the collection of articles analysed.

Three key areas that could benefit from the application of data-driven approaches are the study of the disaggregation of the condition, quality of life, and risk factors and prevalence. One of the greatest challenges for research in these areas regards data curation, particularly its collection and sharing. The study of the course of the condition, including its onset, persistence, and flare-ups, and the design of personalized treatment strategies would be greatly aided by additional longitudinal data. Previous studies have showcased the benefit and need of such data^21,42,43^ and new smartphone apps could facilitate the collection of data outside of a clinical visit. The sharing of data is also crucial, as AD is a complex disease that cannot be fully characterized in a single study. It would be greatly aided by a collaborative system to share and manage data from different studies across the community.

As the development and employment of machine learning and other data-driven approaches gain popularity in healthcare, experts and government agencies have increasingly collaborated to develop guidelines to facilitate the growth of the field and delineate principles of best practice^45,46^. We further underline the need and benefit of cross-disciplinary collaborations for the future of data-driven research on AD and eczema^12^.

## Data Availability

All data and code for the analysis are available at

https://github.com/arianeduverdier/systematic_data-driven_eczema

## Data availability statement

All data and code for the analysis are available at https://github.com/arianeduverdier/systematic_data-driven_eczema.

## Authors’ contributions

AD: Conceptualization, data curation, formal analysis, investigation, methodology, software, validation, visualization, writing-original draft.

AC and RJT: Conceptualization, funding acquisition, project administration, resources, supervision, validation, writing-original draft, writing-review & editing.

## SUPPORTING INFORMATION

Additional Supporting Information may be found in the online version of this article at the publisher’s website:

**Appendix S1**. Supplementary methods.

**Appendix S2**. Additional publication trends.

**Appendix S3**. Atopic dermatitis (AD) and eczema term use.

**Figure S1**. Venn diagram of the number of publications by methodology.

**Figure S2**. Thematic maps generated for each separate methodology and term used.

**Figure S3**. Number of publications in each Latent Dirichlet Allocation (LDA) topic over the years.

**Figure S4**. Hierarchical clustering dendrogram of the eight LDA topics.

**Figure S5**. Keyword co-occurrence network.

**Table S1:** Number of papers published between September 1973 and March 2021 on the application of multivariate statistics (MS), machine learning and artificial intelligence (ML&AI) and Bayesian statistics (BS) to eczema and AD research.

**Table S2**. Top ten most productive countries by the number of publications.

**Table S3**. Summary information on the eight LDA topics.

**Table S4**. Summary table of the five themes and eight topics identified.

