## Supplementary Information for "Data-driven research on eczema: systematic characterization of the field and recommendations for the future"

Running head: PAST AND FUTURE OF DATA-DRIVEN RESEARCH ON ECZEMA

A. Duverdier[1], A. Custovic[2] and R.J. Tanaka[3*]

1. UKRI Centre for Doctoral Training in AI for Healthcare, Department of Computing, Imperial College London, UK
2. National Heart and Lung Institute, Imperial College London, UK
3. Department of Bioengineering, Imperial College London, UK

### Supplementary appendix

**Appendix S1: SUPPLEMENTARY METHODS**

**Literature Search**

All relevant material published on the database, up to March 17th, 2021, was retrieved using six search strings. The keywords “atopic dermatitis” and “eczema” were used separately with each of MS, ML&AI, and BS methodologies. We included both AD and eczema terms, as Frainay et al[1]. demonstrated that the terms are linked to different findings and biased to different disciplines. The terms were used independently to determine if the publications retrieved using one of the two keywords are significantly different to those returned using the other keyword.

The queries for the three methodologies, adapted from the similar analysis conducted by Fontanella et al.[2] on asthma research, are as follows:

- Multivariate statistics (MS): “principal component” OR “discriminant analysis” OR “correspondence analysis” OR “canonical correlation” OR “Markov” OR “factor analysis” OR “factorial analysis” OR “structural equation” OR “latent variable” OR “multidimensional scaling” OR “clustering” OR “latent class” OR “cluster analysis” OR “latent profile” OR “profile regression” OR “mixture model”
- Machine learning and artificial intelligence (ML&AI): “neural networks” OR “deep learning” OR “supervised learning” OR “unsupervised learning” OR “support vector machine” OR “SVM” OR “decision trees” OR “classification trees” OR “regression trees” OR “random forest” OR “machine learning” OR “artificial intelligence” OR “CNN” OR “natural language processing” OR “NLP” OR “reinforcement learning” OR “reinforced learning”
- Bayesian statistics (BS): “Bayesian” OR “Bayes”

The search strings were applied to the title, abstract, and keywords of the publications in the database. The entries returned by the search were manually downloaded in CSV format with all information available for them in the database. All publications without an abstract were removed, as well as conference reviews and erratums.

**Bibliometric Analysis**

The *bibliometrix* R package[3] was used to perform the bibliometric analysis on the collection of documents returned in the search and to visualize the results.

First, the package was used to obtain descriptive statistics of the collection of publications. Descriptive statistics provide key information on the collection of documents, including the most productive countries and authors and the general publication trends.

Second, co-word analysis was applied to produce keyword co-occurrence networks and thematic maps, working under the assumption that the keywords associated to each publication provide an apt overview of the text. Co-word analysis uses networks to represent the knowledge structure of a field where nodes are keywords and edges link keywords that co-occur within the same publication. The size of the nodes and thickness of the edges can be used to represent the frequency of the keywords and the frequency of their co-occurrence within the collection of documents. Community detection algorithms can be used to cluster keyword co-occurrence networks. These resulting clusters can be used to visualize key themes present within a research field and how they are connected to one another. In this study, the keyword co-occurrence networks were clustered using the Louvain community detection algorithm[4], a greedy algorithm that maximizes modularity to find clusters of keywords that maximize intra-cluster connections and minimize inter-cluster connections. The Louvain algorithm was chosen by the authors among the community detection algorithms offered by the package because it has been shown to outperform other community detection algorithms[5] and has been used in similar bibliometric analyses to cluster networks[2,6]; additionally, it gave the most comprehensive clustering (by human judgement) compared to the other algorithms. Both keyword co-occurrence networks and thematic maps were generated using the authors' keywords associated to each publication as they rendered the most informative plots.

Thematic maps are subsequently produced by mapping the themes (clusters from the keyword co-occurrence networks) onto a two-dimensional space using their centrality and density, specifically Callon's centrality and density[7]. As explained in Cobo et al.[8], the centrality is the degree of interaction of the theme with other themes, it measures strength of connections to other themes. More formally, given the equivalence index $e_{ab} = \frac{c_{ab}^2}{c_a c_b}$ which measures the similarity between two keywords where $c_{ab}$ is the number of documents in which two keywords a and b co-occur, and $c_a$ and $c_b$ represent the number in which each one appears. Then, $centrality = 10 \times \sum e_{kh}$ where k is a keyword that belongs to the theme, h is a keyword that is found in other themes, and $e_{kh}$ is the equivalence index. "We can understand this value as a measure of the importance of a theme in the development of the entire research field analyzed"[8]. A high value of centrality means the theme is densely connected to other themes and has therefore likely contributed to the development of the field at large. On the other dimension, density is the strength of the internal connections among the keywords in the theme, it measures the strength of connections within the theme. It can be outlined as $density = 100(\sum e_{ij}/w)$ where i and j are keywords in the theme and $w$ is the total number of keywords in the theme. "This value can be understood as a measure of the theme's development"[8]. Themes are ranked by their relative centrality and density and separated into four quadrants: emerging or declining themes (low centrality and density), niche themes (low centrality and high density), motor themes (high centrality and density), and basic themes (high centrality and low density). Further details on thematic maps can be obtained by reading the paper by Cobo et al.[8].

Refer to the paper by Aria and Cuccurullo[3] for a detailed description of all methods and functions available in the package.

**Probabilistic Topic Modelling**

The Latent Dirichlet Allocation (LDA) algorithm[9] was used to explore the main topics present in the research field. LDA infers latent topic structures from a collection of documents in which each document consists of a collection of tokens. In this study each token represents one word, but the method can be applied to *n*-grams. The order of the words within a document is not considered. The method assumes that each document is composed of a mixture of topics and that each topic consists of a mixture of words (tokens). LDA is a statistical method for estimating both of these distributions: the distribution of topics within each document and the distribution of words within each topic, all of which are Dirichlet distributions. The topic distributions within each document in the collection share a common Dirichlet prior and the word distributions also share a common Dirichlet prior.

More formally[2,9,10], if we have a corpus consisting of $D$ documents, each of which consists of a collection of $N$ words $w_d = \{w_{d,1}, w_{d,2}, \ldots, w_{d,N}\}$ and each word is an item from a vocabulary indexed by $\{1,\ldots,V\}$. And we assume that there are $K$ latent topics with probability distributions $\{\beta_1, \ldots, \beta_K\}$ over the vocabulary. Then, the posterior is

$$p(\beta_{1:K}, \theta_{1:D}, z_{1:D} | w_{1:D}) = \frac{p(\beta_{1:K}, \theta_{1:D}, z_{1:D}, w_{1:D})}{p(w_{1:D})},$$

where $\theta_{1:D}$ are the topic proportions for the $D$ documents ($\theta_{d,k}$ is the topic proportion of topic $k$ in document $d$), $z_{1:D}$ are the topic assignments ($z_{d,n}$ is the topic assignment of word $n$ in document $d$), and $w_{1:D}$ are the observed words ($w_{d,n}$ is word $n$ in document $d$). In this study, the posterior is approximated using Gibbs sampling[11].

The number of topics, $K$, is a key hyperparameter of LDA. A smaller value of $K$ generates broader topics. Increasing $K$ produces more detailed topics, however a value that is too large generates topics that are not semantically coherent. Therefore, it is important to find a value of $K$ such that the topics are at the balance between being detailed and interpretable[10]. There is currently no commonly accepted method for choosing $K$, instead a variation or combination of three main ideas are used[12]: based on domain knowledge (if a set number of topics is expected), human judgement of topic quality (qualitatively assessing the results of models for a range of $K$ values), and topic coherence and exclusivity (calculating the metrics for a range of $K$ values). A combination of all three was used in this study to choose $K$=8 from a considered range of $K$ between two and 16. First, the bibliometric analysis showcased five main themes, thus $K$ had to be greater than five to obtain more knowledge about the topic structure of the research field. Second, a $K$ of eight gave the best balance between sufficiently detailed yet still coherent topics by human judgement of quality. Third, a value of eight gave a good balance between topic coherence and exclusivity, the values of which were plotted over the range of $K$ considered.

Coherence and exclusivity can be calculated for each of the topics in each of the models. Coherence measures whether the top words in a topic tend to co-occur together, since the score is a log probability a value closer to zero indicates that the words in a topic tend to co-occur in the same documents. It is meant to simulate human judgement of topic semantic coherence. Exclusivity measures whether the top words in a topic do not appear as top words in other topics. In this study, as is usually done, only the top 10 words of each topic are considered in the calculation of the metrics. Often, in large datasets of over 1000 documents, a clear peak in coherence can be seen, unfortunately this study only considers 620 documents, so a peak is not as apparent. Instead, a $K$ value can be chosen to balance between topic coherence and exclusivity. We consider both metrics since models with fewer topics have higher topic coherence but lower exclusivity, as models with lower $K$ tend to have topics that are dominated by very common words.

More specifically, topic coherence and exclusivity are calculated using the *topicdoc* R package[13]. Topic coherence is calculated by adding together a score for each pair of top 10 ranked words. This score is the log of the probability that a document containing at least one occurrence of the higher-ranked word also contains at least one occurrence of the lower-ranked word[14,15]. Topic exclusivity is the average, over each of the 10 top words, of the probability of that word in the topic divided by the sum of probabilities of that word in all topics[12,15].

**Commonly applied analytic methods**

For MS and ML&AI publications, search strings representing the methods were searched in the title, abstract, and keywords of each publication to obtain the number of publications they appear in. For BS, a manual inspection of the articles was performed to find the methods used in each publication.

**Appendix S2: Additional publication trends**

A median of two articles were published per year from 1973 to 2002, 14 for 2003 to 2012, and 42 for 2013 to 2019. In 2020 alone, 105 papers were published; and 2.5 months into 2021, 21 papers were already published. 37 articles applied two of the three methodologies and two articles applied all three methodologies (Figure S1). The greatest overlap occurred between MS and ML&AI, with 24 articles in common. Price's model[16] theorizes that the growth of a scientific field undergoes three key phases: first, an initial stage characterized by growth in small increments; second, a stage of exponential growth in research as the field draws in an increasing number of researchers; third, the field approaches saturation[10,17]. Following this model, it is likely the application of MS, ML&AI, and BS to AD and eczema research has recently entered the second phase, showing that the field still has much growth and development to undergo.

The publications retrieved included a total of 3,973 authors, 17 of which produced single-authored publications and the remaining authors participated in multi-authored publications. The search returned an average of 0.16 documents per author, 6.41 authors per publication and a collaboration index of 6.57. The average number of citations per publication was 27.69. The USA, the UK, Germany, and France are the four most productive countries, measured by the number of publications (94, 61, 40, and 39 respectively) and total citations (4,710, 2,020, 1,642, and 1,063 respectively) (Table S2). Each of the top ten most productive countries collaborated with other countries (Table S2). Germany has the highest percentage of multiple country publications (55%, 22 of 40 publications) and the USA has the largest number of multiple country publications (26 publications). The ten most productive countries, measured in number of publications, are located in Europe, Asia, North America, and Australia; each of them has participated in multiple country publications. This showcases the collaborative nature of the field and links to the worldwide occurrence of AD and eczema.

The query used to retrieve eczema papers included atopic eczema (AE). 35 papers in the collection are tagged with the term AE, four of which only use the term AE, while the remaining 31 also include AD and/or eczema.

It is also important to note that many of the papers returned in the search are not specific to AD and eczema, but instead study allergy and associated allergic conditions, including AD/eczema, food allergy, asthma, and allergic rhinitis. This indicates the current interest in studying the relationship between allergic conditions and the continued need to study AD and eczema more specifically.

**Appendix S3: Atopic dermatitis and eczema term use**

The continued absence of a consensus in nomenclature has resulted in the co-existence of two main terms for the skin condition, AD and eczema. Within the literature, the majority of publications from 1945-to-present are annotated with one of the two terms, but not both[1,18]. This separate use of terms leads to several challenges including confusion in drug development, biomarker discovery, research in epidemiology, and public health and reimbursement, as well as an impact on data mining[1,19]. Frainay et al[1]. demonstrated the impact of the co-existence of the two terms on the retrieval of information, finding that under five percent of the total AD and eczema articles in PubMed published between 1945 and 2017 are annotated with both terms. The literature search conducted in this paper reflects this general trend, with only approximately 16% of the articles (100 of 620 articles) jointly annotated with eczema and AD terms. Although the terms are often seen as synonymous and used interchangeably, two recent studies have shown that term use varies between different fields of study, supporting the idea that the publications and findings associated to each term are not homogeneous. Frainay et al.[1] demonstrated that the terms are linked to different findings and biased to different disciplines. For instance, the AD term was linked to more publications related to veterinary science, biochemistry, and cellular and molecular biology, while the eczema term was associated to infectious disease, public health, and the respiratory system. In this study, the five general themes of research are found in both AD and eczema literature, however, the term eczema is often used in studies that regard its association to other allergic or skin diseases, while AD is used more often in studies on its treatment and quality of life. Additionally, term use is not uniform within the methodologies. Studies employing ML&AI tend to use the eczema term more heavily, while separate methods within MS and BS see varying biases in term use. For instance, within the MS methodology, factor analysis is used by a ratio of approximately 1.8 to one in favor of AD, but discriminant analysis has a ratio of two to one in favor of eczema. This reflects the previously proposed notion that AD and eczema term use may be associated to different research communities who have different views on nomenclature.

#### SUPPLEMENTARY FIGURES

**Figure S1**: Venn diagram of the number of publications by methodology.

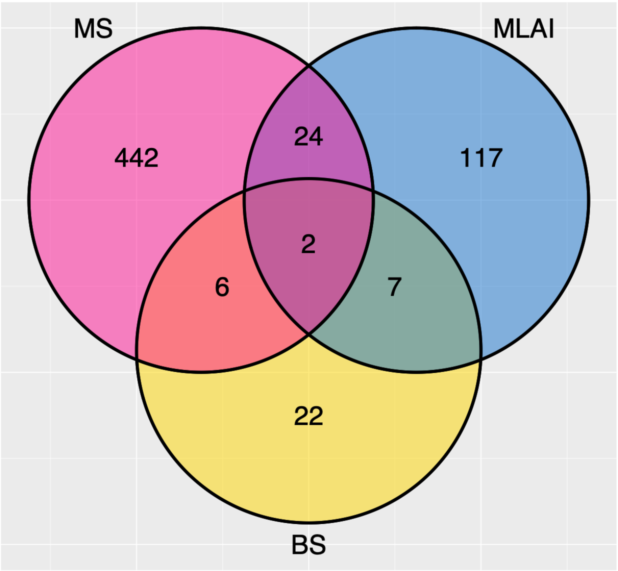

**Figure S2**: Thematic maps of the application of multivariate statistics (MS), machine learning and artificial intelligence (ML&AI), and Bayesian statistics (BS) to eczema and atopic dermatitis research. Generated for each separate methodology, **(A)** MS, **(B)** ML&AI, and **(C)** BS**.** And the term used, **(D)** AD and **(E)** eczema. Plots were created using the top 100 authors' keywords. Themes are separated according to centrality (the degree of interaction of the theme with other networks) and density (the strength of internal connections among keywords in the theme). Up to six of the most significant keywords in the associated theme are labelled.

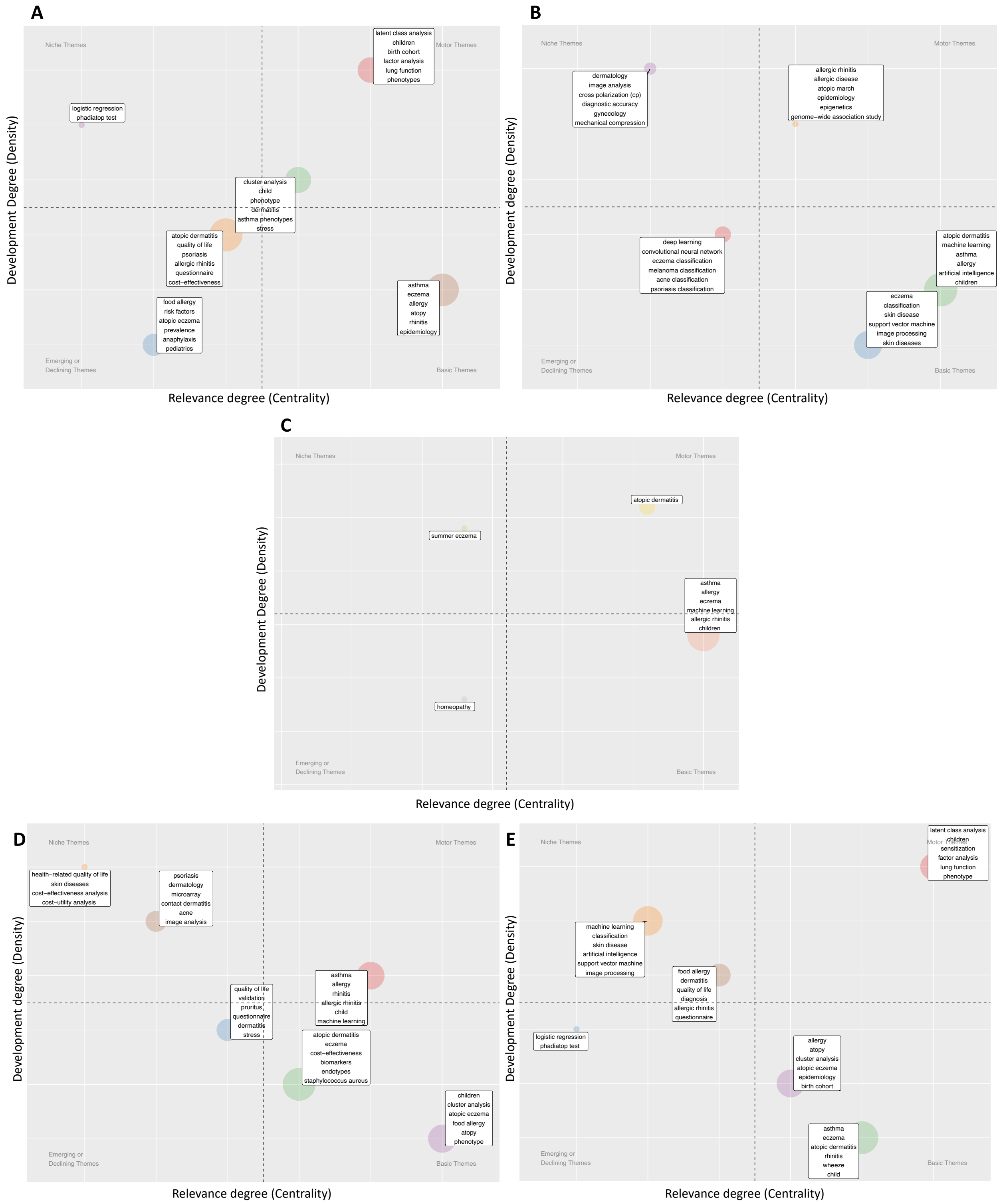

**Figure S3**: Number of publications in each LDA topic over the years. Each publication is assigned its top three most probable topics, provided the probabilities are greater than 0.1.

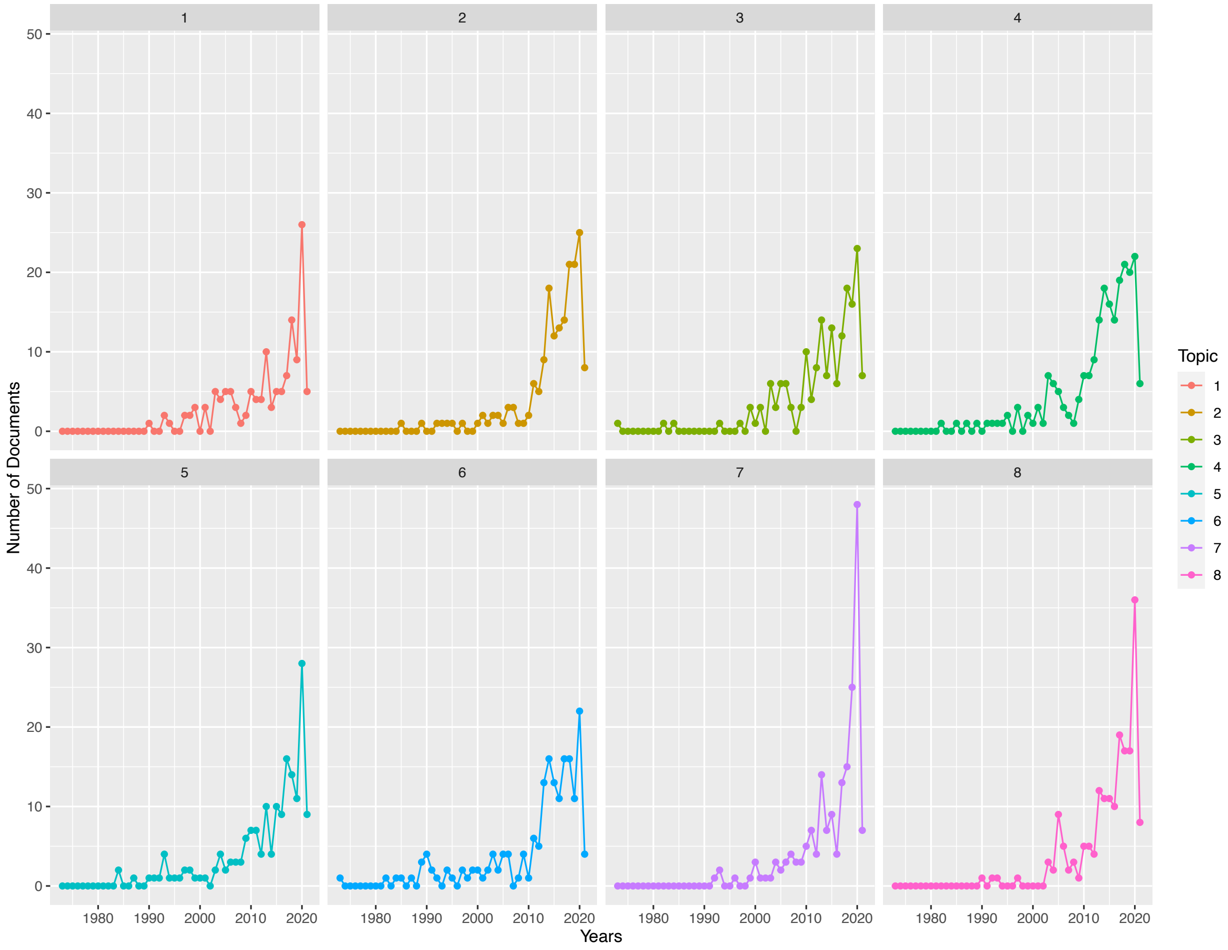

**Figure S4:** Hierarchical clustering dendrogram of the eight LDA topics. Y-axis: distance between the clusters (topics) as computed with Hellinger distance.

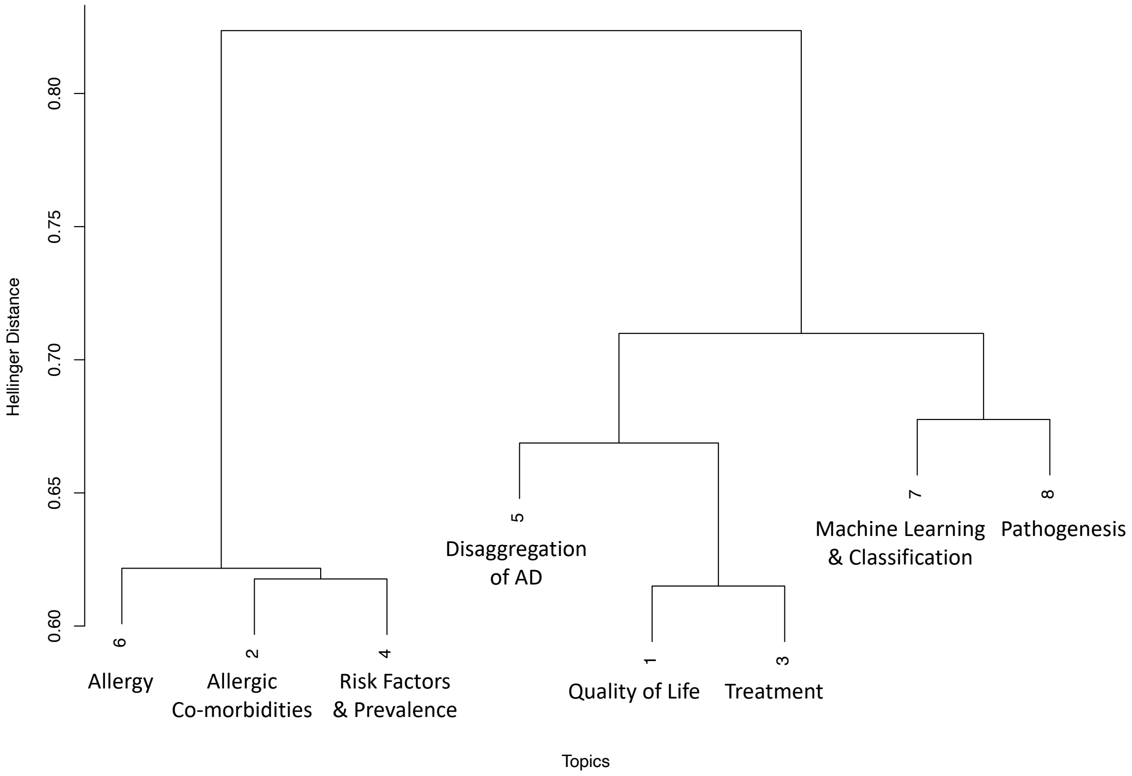

**Figure S5**: Keyword co-occurrence network of the full collection of publications. Generated using the top 50 authors' keywords. Nodes represent authors' keywords and edges connect keywords that co-occur in a document. The thickness of the edge corresponds to the number of times these keywords co-occur. The size of the node is proportional to the strength of the keyword. The Louvain community detection algorithm is used to cluster the network.

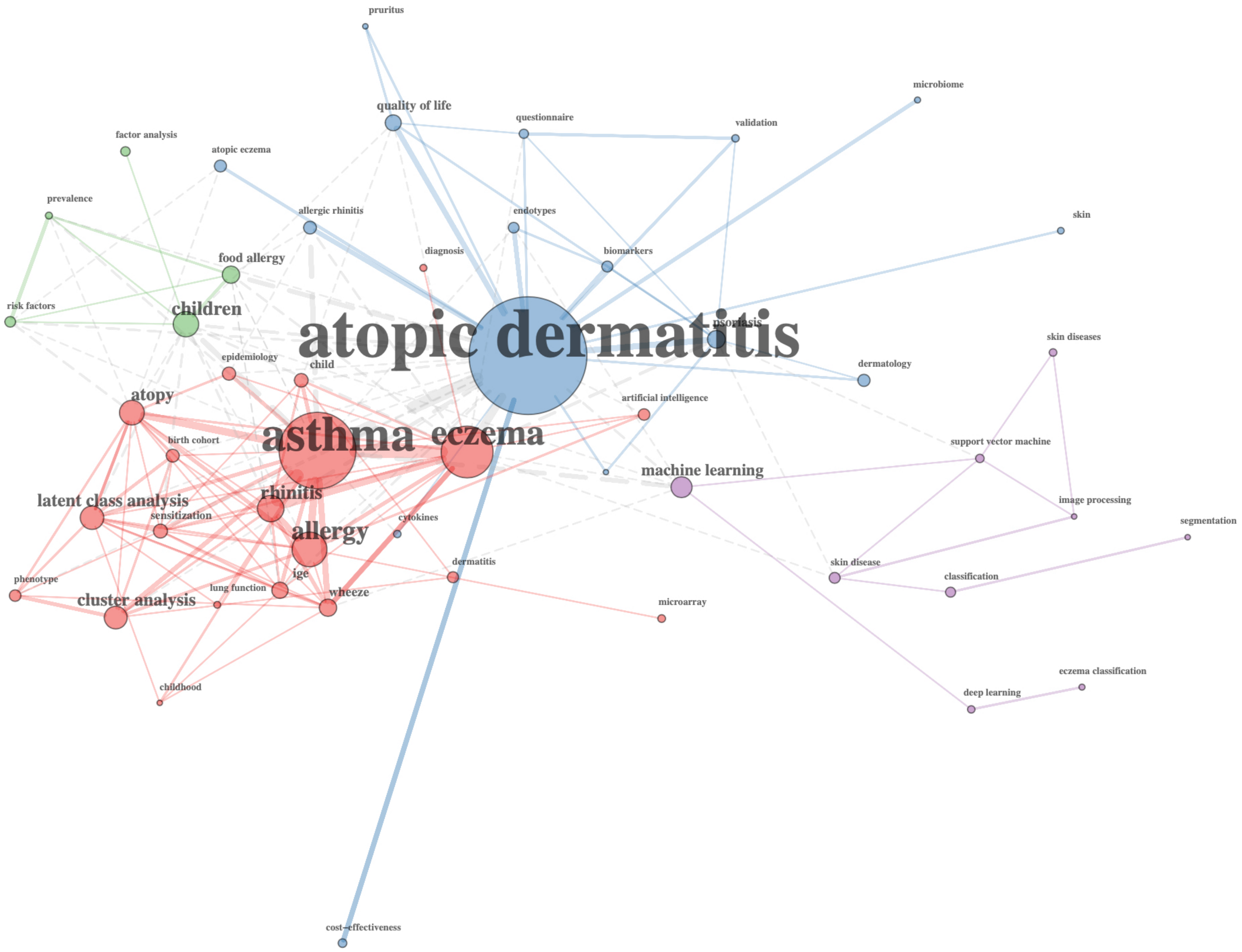

#### SUPPLEMENTARY TABLES

**Table S1**: Number of papers published between September 1973 and March 2021 on the application of multivariate statistics (MS), machine learning and artificial intelligence (ML&AI) and Bayesian statistics (BS) to eczema and atopic dermatitis (AD) research. Rows indicate term(s) used in publication, only AD, only eczema, or both.

| | MS | ML&AI | BS |
|---|---|---|---|
| AD | 212 | 53 | 11 |
| Eczema | 182 | 78 | 10 |
| Both | 80 | 19 | 6 |
| TOTAL | 474 | 150 | 37 |

**Table S2**: Top ten most productive countries, by the number of publications. Single country publications (SCP) and multiple country publications (MCP).

| | Documents Published | | | | Citations | | |
|---|---|---|---|---|---|---|---|
| Country | Rank by Total Document s | Total | SCP | MCP | Rank by Total Citations | Total | Average Document |
| USA | 1 | 94 | 68 | 26 | 1 | 4710 | 50.11 |
| UK | 2 | 61 | 40 | 21 | 2 | 2020 | 33.11 |
| Germany | 3 | 40 | 18 | 22 | 3 | 1642 | 41.05 |
| France | 4 | 39 | 31 | 8 | 4 | 1063 | 27.26 |
| China | 5 | 27 | 24 | 3 | 13 | 275 | 10.19 |
| Korea | 6 | 27 | 22 | 5 | 12 | 291 | 10.78 |
| Japan | 7 | 26 | 22 | 4 | 6 | 506 | 19.46 |
| Sweden | 8 | 21 | 13 | 8 | 5 | 627 | 29.86 |
| Denmark | 9 | 19 | 16 | 3 | 7 | 449 | 23.63 |
| Australia | 10 | 17 | 6 | 11 | 8 | 439 | 25.82 |

**Table S3**: Summary information on the eight LDA topics. Topic proportion is the mean probability of the topic over all documents.

| Topic | Topic Description | Proportion | Topic Size (Number of Words) | Number of Documents Topic Appears | Number of Documents Topic is Most Probable |
|---|---|---|---|---|---|
| 1 | Quality of Life | 0.118 | 151 | 97 | 71 |
| 2 | Allergic Co-morbidities | 0.114 | 111 | 125 | 69 |
| 3 | Treatment | 0.127 | 182 | 112 | 67 |
| 4 | Risk factors & Prevalence | 0.140 | 158 | 157 | 100 |
| 5 | Disaggregation of AD | 0.102 | 126 | 104 | 51 |
| 6 | Allergy | 0.119 | 110 | 127 | 70 |
| 7 | Machine Learning & Classification | 0.155 | 181 | 141 | 104 |
| 8 | Pathogenesis | 0.126 | 164 | 136 | 87 |

**Table S4**: Summary table of the five themes and eight topics identified.
Themes were determined via a bibliometric analysis, specifically through plotting a thematic map of the collection of documents. The themes were ranked from 1 to 5 according to their relative density and centrality. The topics obtained from applying Latent Dirichlet Allocation (LDA) to the collection of documents were then manually allocated to the theme that best corresponds. The topic proportions (mean probability of the topic over all documents in the collection) were summed for themes that contained more than one topic. Each document in the collection is assigned the LDA topic that is most probable, these numbers are summed for themes that contain more than one topic to return the total number of documents theme is most probable. The number of documents the theme is most probable is then separated according to methodology, multivariate statistics (MS), machine learning and artificial intelligence (ML&AI), or Bayesian statistics (BS) that is applied in the document. Note that some documents apply more than one methodology and are therefore counted multiple times. According to these numbers the most and least popular methodology is selected for each theme.

| Theme | Theme 1 | Theme 2 | Theme 3 | Theme 4 | Theme 5 |
|---|---|---|---|---|---|
| Title | Image analysis and classification | Allergic co-morbidities | Disaggregation of AD | Quality of life | Risk factors and prevalence |
| Density (development) | 2 | 1 | 3 | 4 | 5 |
| Centrality (relevance) | 5 | 1 | 3 | 2 | 4 |
| Type of Theme | Niche | Motor | - | Basic | Emerging |
| LDA Topic Numbers | 7 | 2, 6 | 5, 8 | 1, 3 | 4 |
| LDA Topic Titles | Machine learning and classification | Allergic co-morbidities, Allergy | Disaggregation of AD, Pathogenesis | Quality of life, Treatment | Risk factors and prevalence |
| Total proportions of LDA topics | 0.155 | 0.233 | 0.228 | 0.245 | 0.140 |
| Total number of documents theme is most probable | 104 | 139 | 138 | 138 | 100 |
| Number of MS documents | 28 | 125 | 115 | 119 | 86 |
| Number of ML&AI documents | 82 | 24 | 23 | 13 | 8 |
| Number of BS documents | 4 | 11 | 2 | 11 | 9 |
| Most popular methodology | ML&AI | MS | MS | MS | MS |
| Least popular methodology | BS | BS | BS | BS | ML&AI |
